# From Sparse Data to Smart Decisions: Region-Specific Policy Evaluation via Simulation

**DOI:** 10.64898/2025.12.05.25341712

**Authors:** Carson Dudley, Daniel Bergman, Harsh Jain, Kerri-Ann Norton, Erica Rutter, Marisa Eisenberg, Trachette Jackson

## Abstract

Effective infectious disease response requires region-specific guidance, because the contact structures that drive local transmission and shape intervention effects vary from community to community. Agent-based models (ABMs) represent these structures and the individual heterogeneity underlying them, which makes them well suited to local intervention analysis, but they are computationally expensive and fitting their parameters to data is difficult. We present a framework that avoids direct ABM fitting by using a surrogate ordinary differential equation (ODE) model as an intermediary between routine surveillance data and a mechanistically rich ABM. Such data rarely identify ABM parameters uniquely: many combinations of transmissibility, severity, and reporting can reproduce the same observed trajectories. Rather than committing to a single best-fit configuration, the framework retains the full set of ABM parameter combinations consistent with observed case and hospitalization data and propagates this uncertainty into every downstream intervention estimate. We apply the framework to COVID-19 in nine demographically distinct Michigan counties as an illustrative case study. For each county, we recover the ABM parameter regions consistent with local case and hospitalization data, verify that the calibrated model reproduces the observed trajectories, and then evaluate four non-pharmaceutical interventions across those regions. We find that the ranking and robustness of the interventions differ substantially across counties, and that simple demographic predictors do not reliably predict which will succeed. We use intervention implementation assumptions (e.g., for compliance and efficacy) drawn from COVID-19 literature, but the framework is general and can take user-specified assumptions and show what conclusions they would lead to. By coupling mechanistic ABMs with explicit uncertainty propagation, the approach provides a generic route to local, uncertainty-aware intervention analysis for COVID-19, influenza, measles, and other pathogens.

## 1 Introduction

Local health agencies routinely face decisions about which non-pharmaceutical interventions (NPIs) to use during respiratory disease outbreaks. Determining the best intervention requires region-specific modeling tools, because demographics, comorbidities, contact patterns, and healthcare capacity vary substantially across communities and drive real-world outcomes [1, 2]. National or global parameter estimates rarely capture this variation [3]. Without locally-grounded analyses, resource allocation and intervention planning risk being poorly matched to actual needs [4].

A common approach is to fit simplified ordinary differential equation (ODE) models, such as SIR-type compartmental models, to surveillance data [5, 6]. These models are computationally efficient and analytically tractable. They are well-suited to rapid assessment and forecasting. However, they typically assume random mixing across the population. This makes it difficult to evaluate interventions that act on specific contact networks, such as school closures or workplace restrictions, because the relevant network structure is not represented. Agent-based models (ABMs) offer a complementary approach [7, 8]. They simulate individuals and their interactions in homes, schools, workplaces, and community settings. This makes them well-suited to evaluating interventions that target specific contact structures. ABMs are computationally more expensive than ODEs, and parameter inference for ABMs requires care [9]. A range of methods have been developed for this purpose, including approximate Bayesian computation, gradient-based methods [10], and others.

We use a surrogate-informed inference approach to fit ABMs based on the SMoRe ParS framework (Surrogate Modeling for Reconstructing Parameter Surfaces) [11, 12]. SMoRe ParS uses a simplified ordinary differential equation (ODE) model as an intermediary between observed data and the more complex ABM. The same ODE surrogate is fit to real surveillance data and to synthetic outputs from the ABM across many parameter combinations. Comparing the two sets of fits identifies which ABM configurations produce dynamics consistent with observed case and hospitalization trajectories. We use SMoRe ParS because it has been validated in other multi-scale settings where agent-based simulations are available as well as mean-field ODE descriptions [11, 12]. Other ABM inference methods could in principle be substituted, our contributions do not depend on this specific choice.

The most important aspect of our methodology is how it handles parameter uncertainty. Routine case and hospitalization data do not uniquely identify individual ABM parameters. Many combinations of transmissibility, latent period, asymptomatic fraction, reporting rate, and severity can reproduce the same observed trajectories. Rather than select a single best-fit configuration and treat it as the truth, we retain the full set of ABM parameter combinations consistent with the data. We then propagate this set forward through intervention simulations. The result is a distribution of intervention outcomes that reflects the underlying uncertainty in transmission dynamics, severity, and reporting. This distinguishes interventions that are robust across the data-consistent parameter regions from those whose effectiveness depends sensitively on parameters that the data cannot fully resolve. The intervention assumptions in any analysis, including compliance rates and the contact-network effects of each intervention, are also uncertain. We use values drawn from the literature (section 4), but our framework is general, so users can substitute their own assumptions and re-run the analysis without changes to the inference pipeline.

We demonstrate the framework through a case study of COVID-19 in nine demographically distinct Michigan counties during late 2020. We fit the surrogate model jointly to county-level case and hospitalization time series and identify the corresponding regions of ABM parameter space. We then evaluate four NPIs across these regions: school closures, stay-home-if-symptomatic policies, non-essential workplace closures, and general social distancing. A schematic overview of the framework is shown in Figure 1. The ranking and robustness of NPIs differ substantially across counties, and simple demographic predictors such as age structure, population density, or workforce participation do not reliably predict which interventions will be most effective. School closure efficacy tracks child population as would be expected, but the other interventions do not align with their most natural demographic proxies (e.g., workplace closures and labor force participation rate). This motivates mechanistic, region-specific evaluation rather than reliance on demographic heuristics. Our framework provides one such route, and can be generalized to other pathogens and to settings where routine surveillance data is the main available signal.

**Figure 1:**
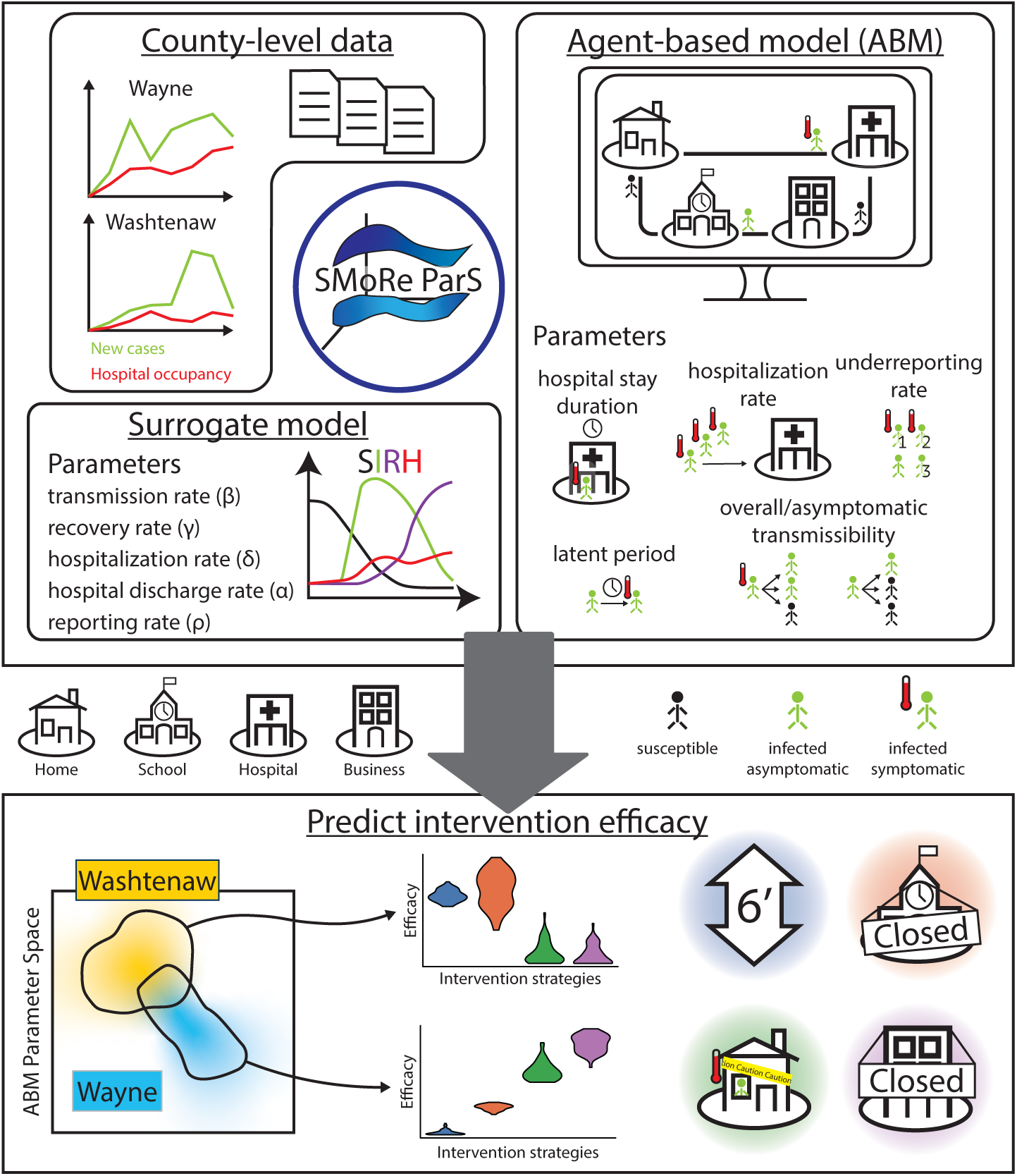
Schematic illustrating how SMoRe ParS integrates county-level data with an agent-based model and surrogate model to make predictions about specific interventions within counties. The four interventions we evaluate are social distancing (blue), school closure (red), stay-home-if-symptomatic (green), and non-essential work-closure (purple).

## 2 Results

### ABM Reproduces Observed Epidemic Dynamics

To simulate county-level outbreak dynamics under real-world conditions, we used an agent-based model (ABM) implemented in the Framework for Reconstructing Epidemiological Dynamics (FRED) simulation platform (see Methods: Agent-Based Simulations) [13]. The model explicitly represents individuals and their interactions across home, school, work, and community settings, capturing demographic and behavioral heterogeneity relevant to local transmission dynamics. We focused our main analysis on three demographically distinct Michigan counties: Washtenaw County (younger, university-centered), Wayne County (densely populated urban center with high comorbidity burden), and Ontonagon County (rural with an aging population and limited local healthcare capacity). These counties were selected to span a wide range of demographic and infrastructural settings. To evaluate whether key conclusions, such as the relative effectiveness of specific interventions, held consistently within similar contexts, we also applied the framework to six additional counties approximately matched on structural features (e.g., urbanicity, age distribution, and healthcare access). Results for these are included in the Supplementary Information.

Using the SMoRe ParS framework, we first calibrated a simplified, ODE-based, SIR-type surrogate model to observed COVID-19 case and hospitalization data on a county-by-county basis [14]. We then pulled back these data-constrained surrogate fits to get corresponding county-specific data-consistent ABM parameter regions. This surrogate-informed mapping enabled us to efficiently find data-consistent parameter sets without directly fitting the full ABM to data.

Figure 2 displays the results of calibrated ABM simulations in the three representative Michigan counties, obtained by averaging ten stochastic runs using parameter vectors sampled from the surrogate-constrained regions. The simulated epidemic time-courses (blue lines) closely track the reported case trajectories (black dots), accurately capturing both the rise and fall of the COVID-19 wave from late August to December 2020. This fit demonstrates that the surrogate-guided inference approach can successfully recover ABM configurations that explain observed dynamics without overfitting transient noise. We further validate our ABM fits by assessing the fit on a held-out data window in the appendix. The inferred parameter ranges for each county are reported in the appendix.

**Figure 2:**
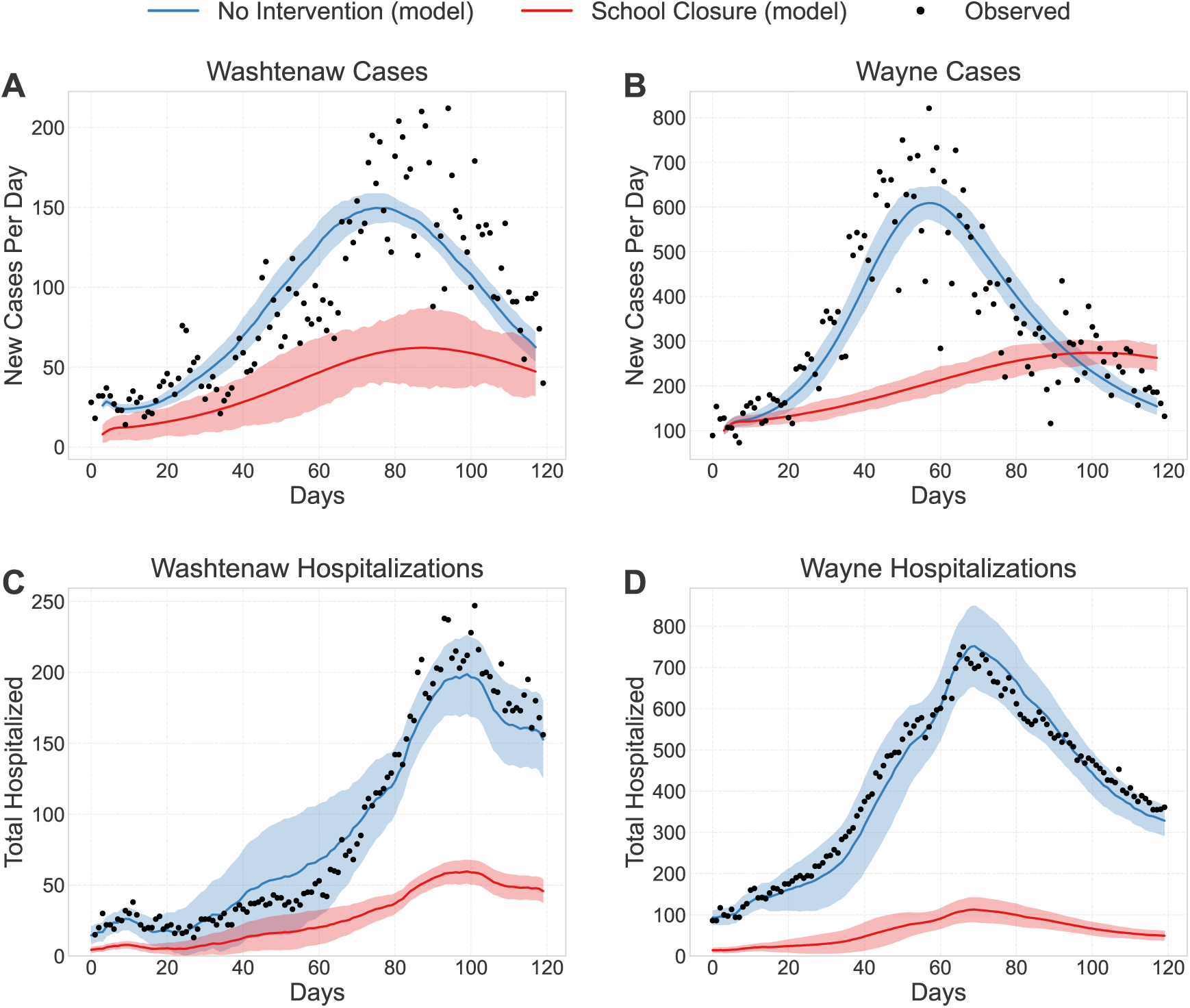
Calibration of the agent-based model (ABM) to COVID-19 cases (top row) and hospitalizations (bottom row) in Washtenaw (left) and Wayne (right) counties. Black dots represent observed data, with values fewer than 5 censored to protect patient confidentiality. Blue curves show ABM output averaged across ten simulation runs with no interventions, and red curves show ABM output under a school closure scenario. Shaded areas represent ±1 SD across simulations with parameters varied within the confidence regions. Ontonagon County is not shown because case and hospitalization counts were so low that nearly all values were censored, particularly for hospitalizations.

### Translating Uncertainty into Intervention Analysis

For decision-making, the critical challenge is not just estimating parameters, but understanding how uncertainties and trade-offs between them shape policy outcomes. Our framework demonstrates these trade-offs by revealing the structure of parameter dependencies that arise from the calibration process. For example, the observed case trajectory can be reproduced by balancing force of infection against case reporting rate: higher transmission with lower detection yields similar observed cases versus lower transmission with higher detection. By propagating this uncertainty into intervention evaluation, our framework reflects data-driven uncertainty.

To visualize these dependencies, we applied kernel density estimation to the top-fitting parameter sets from the surrogate model. Figure 3 displays contour plots for four surrogate model parameter relationships: (A) surrogate case reporting rate vs. force of infection, (B) hospitalization rate vs. discharge rate, (C) hospitalization rate vs. surrogate hospitalization reporting rate, and (D) basic reproduction number (a measure of transmissibility) vs. surrogate case reporting rate. The strength of parameter dependencies varies. Panels A and D have ridge-like structures indicating compensatory behavior: higher transmissibility can be offset by lower reporting rates to produce similar observed case trajectories. Panels B and C show weaker correlations, suggesting these parameter pairs are more independently constrained by the data.

**Figure 3:**
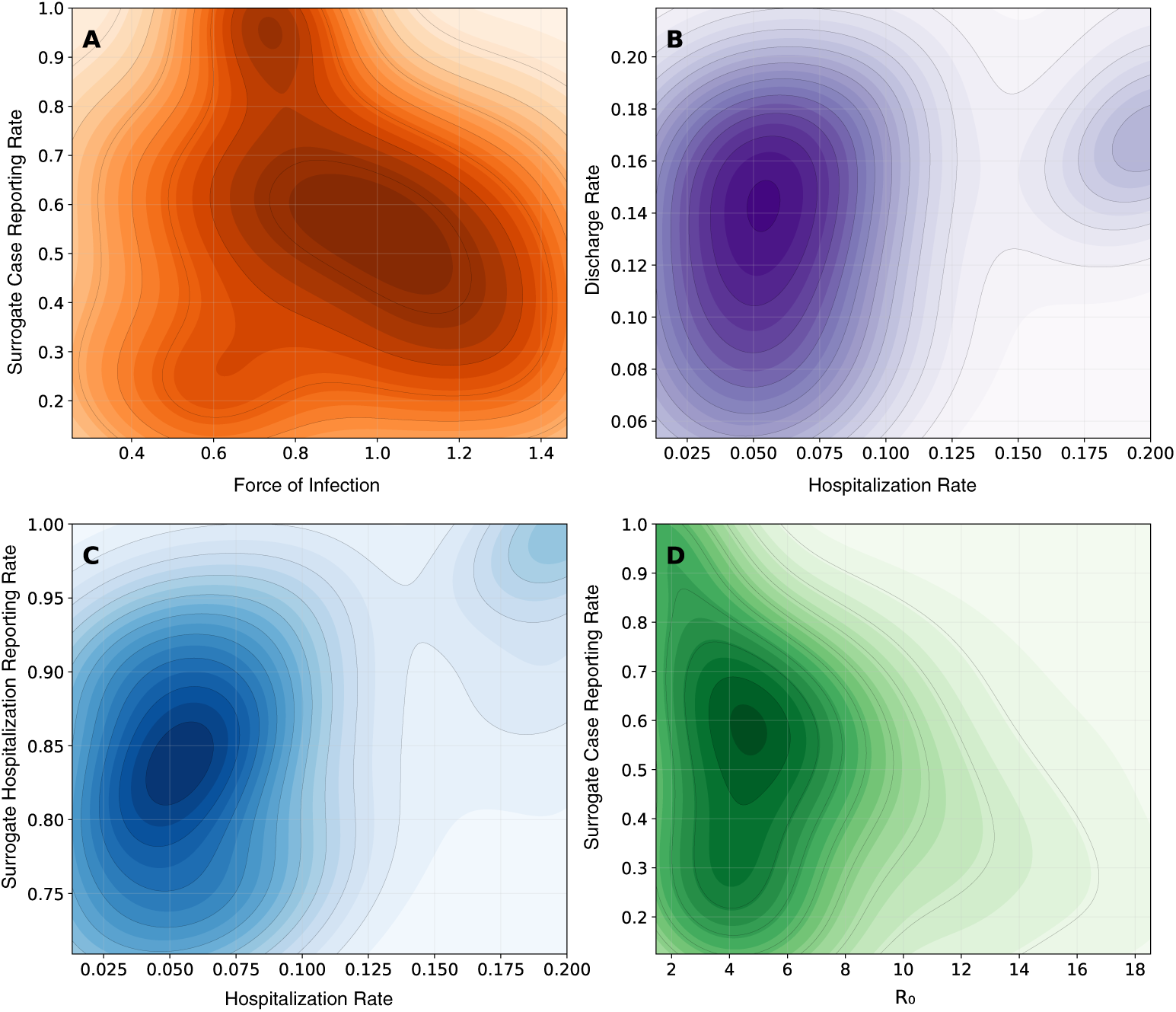
Parameter dependencies in the surrogate model. Contour plots show kernel density estimates over the top-fitting parameter sets for Washtenaw County. (A) Force of infection vs. surrogate case reporting rate shows a clear compensatory trade-off, where higher transmission can be offset by lower reporting. (B) Hospitalization probability vs. discharge rate (1/duration). (C) Surrogate hospitalization rate vs. surrogate hospitalization reporting rate. (D) Basic reproduction number (*R*_0_) vs. surrogate case reporting rate also exhibits a negative relationship. The strength of these dependencies varies across parameter pairs, reflecting different levels of identifiability from the observed data. These parameter uncertainties and correlations are explicitly propagated in downstream intervention analyses.

The data strongly constrain some parameter combinations with weaker identifiability for others. In addition to data fitting, parameter tradeoffs can impact intervention efficacy analysis. Parameters that are not very compensatory for reproducing the observed dynamics may still interact in determining intervention effects. Visualizing the full structure of parameter dependencies underscores why point estimates are insufficient and motivates propagating complete parameter distributions into intervention simulations.

### Robustness of Interventions under Uncertainty

Having calibrated the models for each county, we next evaluated the robustness of candidate non-pharmaceutical interventions (NPIs) by simulating each intervention across the full distribution of ABM parameter combinations identified as consistent with local surveillance data. The interventions considered included school closure, symptomatic isolation messaging (“stay home if sick”), non-essential workplace closure, and general social distancing. The specific implementations for each intervention were drawn from the literature (section 4). These choices are illustrative and could be substituted with alternative assumptions without changes to the inference pipeline.

To reflect the range of epidemic conditions a decision-maker might face, we sampled from the set of ABM parameter combinations whose surrogate projections fell within the 95% confidence region derived from the observed data. Then, we applied each intervention to these simulated outbreaks. This approach allowed us to capture variation in intervention effectiveness arising from uncertainty in transmission dynamics, disease severity, and underreporting.

Figure 4a-d summarizes the distribution of reductions in peak hospitalizations achieved by each intervention across counties and parameter regimes. Each violin represents the range of outcomes generated by varying underlying ABM parameters within the data-consistent region, illustrating both the median effectiveness and the uncertainty associated with each strategy. Figure 4f summarizes robustness, measured as the interquartile range (IQR) of peak reductions in hospitalizations. Interventions with smaller IQRs are more consistent across uncertainty, while larger IQRs reflect greater variability.

**Figure 4:**
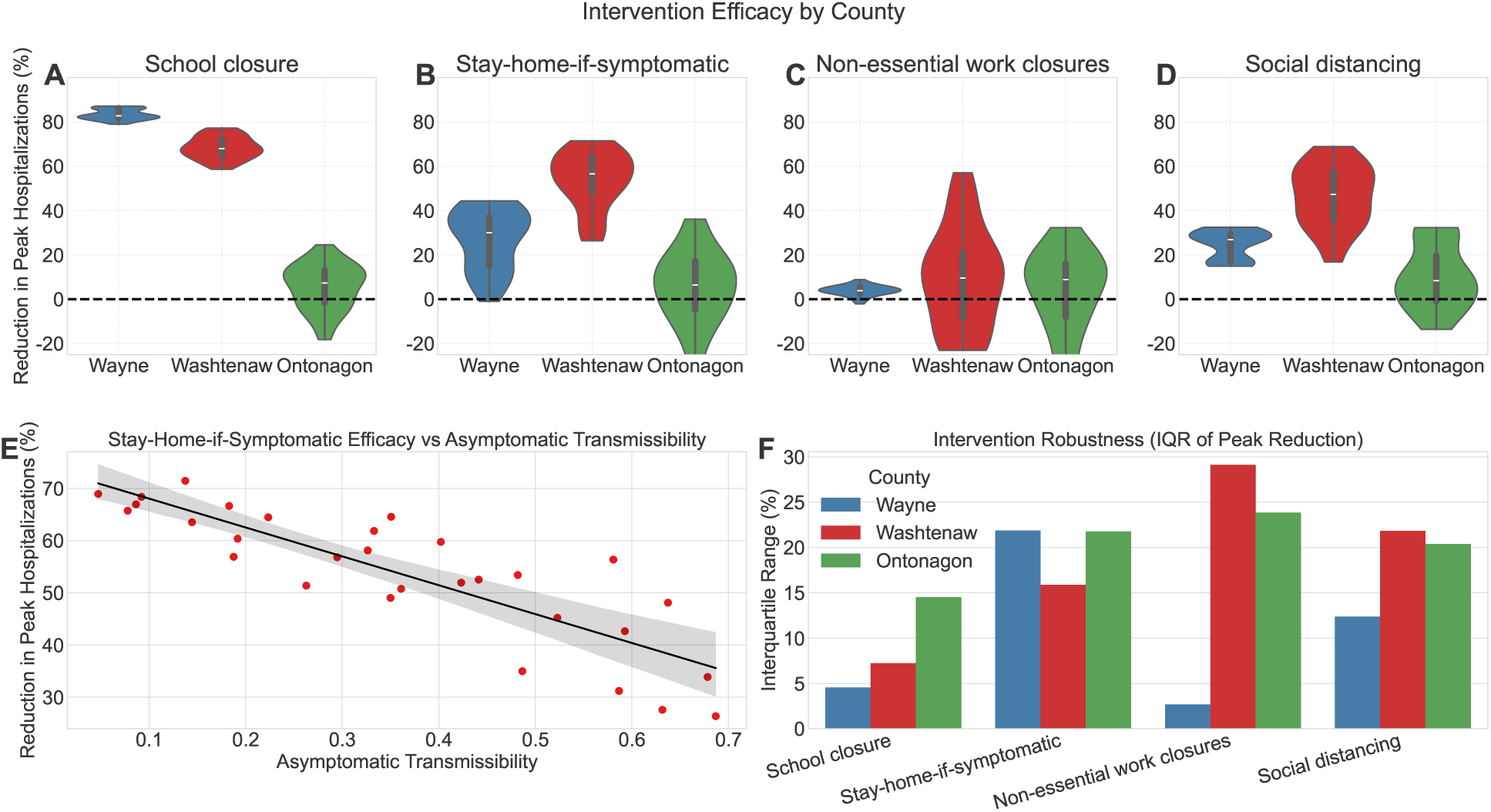
Intervention effectiveness across counties under uncertainty. Each panel shows the distribution of reductions in peak hospitalizations (%) for a single intervention, simulated across all data-consistent ABM parameter combinations in three Michigan counties. **(A)** K-12 school closures produced the largest and most consistent reductions in peak burden in Wayne and Washtenaw Counties (medians: ∼85% and ∼70%, respectively), with more modest and variable effects in Ontonagon (∼10%). **(B)** Stay-home-if-symptomatic policies were effective in Washtenaw (median ∼60%) but showed reduced and more variable impact, reflecting sensitivity to asymptomatic transmission. **(C)**Non-essential work closures yielded modest and inconsistent effects in all counties, with median reductions of 5-10% and high variance in Washtenaw and Ontonagon counties. **(D)** General social distancing produced moderate reductions in Washtenaw and Wayne, but limited and highly variable impact in Ontonagon. Shaded violins represent the distribution of outcomes across parameter regimes; negative values reflect stochastic variability in low-effect-size scenarios. **(E)** Effectiveness of stay-home-if-symptomatic interventions declines as the asymptomatic proportion increases, as shown for Washtenaw County. Each point represents a simulation under a different asymptomatic transmission assumption, and the trend highlights the structural vulnerability of symptom-based isolation to undetected spread. **(F)** Robustness of each intervention, measured as the interquartile range (IQR) of peak hospitalization reductions across retained parameter sets. Lower IQR indicates more stable performance across uncertainty. School closures consistently show low IQRs (high robustness), while symptom-based isolation and social distancing exhibit greater variance.

#### School Closure Impact Varies with Local Age Distribution

Under our implementation, which fully eliminates school and classroom contact rates, school closures produced large and stable reductions in peak hospitalizations in Wayne and Washtenaw Counties (figure 4a), with median reductions of approximately 85% and 70%, respectively. The effect was stable across the data-consistent parameter region, reflecting the role that school-based contact networks play in transmission in counties with substantial student populations and higher overall population density. Empirical analyses of statewide school closures during the first wave of COVID-19 in the United States estimated weekly reductions of approximately 62% in incidence, with larger reductions when closures were implemented earlier [15], so it is not surprising that counties with high density of students see reduction magnitudes slightly higher than this when school closures are implemented at the beginning of the wave.

In contrast, Ontonagon County saw much smaller and more variable reductions, with a median effect around 8% and wide stochastic spread. This result is consistent with expectations: Ontonagon has an aging population and relatively few school-aged children, so school closures remove a smaller share of transmission opportunities. In such low-density, older populations, the impact of school-based interventions is structurally constrained.

These patterns highlight the importance of tailoring intervention strategies to local demographic and contact structures. Where schools serve as major hubs of transmission, school closures can be a highly effective and reliable early intervention for slowing initial outbreak growth or flattening epidemic peaks. However, it is important to acknowledge that prolonged school closures may not be practical or sustainable throughout an extended outbreak or pandemic, given their impacts on education, childcare, and family economic stability. School closures also tend to be more socially and politically contentious than other interventions, requiring consideration of community values and trade-offs. In older or rural populations where schools contribute less to overall transmission, other strategies may be more impactful and face fewer implementation barriers.

#### Symptom-Based Interventions Are Vulnerable to Asymptomatic Spread

Stay-home-if-symptomatic policies reduced peak burden in many scenarios, but their effectiveness varied substantially with the proportion of asymptomatic infections (figure 4b). In Washtenaw County, peak hospitalization reductions ranged from approximately 18% to 68% depending on asymptomatic spread assumptions, while Ontonagon County saw even wider variation and lower median effectiveness. The failure to capture asymptomatic transmitters limits this intervention’s robustness under uncertainty. Policymakers should exercise caution when relying on symptom-based isolation alone, as performance can degrade sharply if asymptomatic transmission is more prevalent than anticipated (figure 4e).

#### Workplace Closures Have Modest and Variable Effects Across Settings

Non-essential work closures led to modest reductions in peak hospitalizations across counties (figure 4c). In Wayne County, the intervention produced consistent but small benefits, with peak reductions centered around 4%. In Washtenaw and Ontonagon Counties, outcomes were more variable, spanning from moderate reductions to small apparent increases in peak burden (negative y-values in figure 4a-d).

These occasional negative estimates do not indicate that workplace closures were harmful; rather, they reflect the combination of small overall effect size and stochastic variability inherent in agent-based simulations. When effects are modest, random variation in outbreak timing or seeding can dominate the signal in individual runs.

The relative lack of variability in predicted hospitalization reductions in Wayne County likely reflects its large population size, which buffers against stochastic fluctuations and yields smoother aggregate dynamics. In smaller or more heterogeneous settings, such as Washtenaw and Ontonagon, workplace closures yield more variable outcomes, despite using the same intervention assumptions. Overall, workplace closures may contribute marginally to outbreak mitigation, but are unlikely to serve as reliable standalone interventions, especially in settings where work-based contacts do not dominate transmission.

#### General Distancing Is Highly Sensitive to Transmission Dynamics

General social distancing produced variable reductions in peak hospitalizations across counties and parameter regimes (figure 4d). Median effects ranged from 8–47%, with the highest reductions seen in urban counties like Wayne. In contrast, reductions were smaller and more variable in Ontonagon, where overall transmission is lower and social mixing is more limited.

Importantly, compliance assumptions were held constant across simulations. The variability in outcomes instead reflects sensitivity to underlying transmission characteristics—such as asymptomatic transmissibility and baseline contact rates. When asymptomatic spread is high, for example, social distancing among symptomatic individuals alone may fail to contain transmission.

While general distancing can meaningfully reduce burden under favorable conditions, its impact is highly parameter-dependent. It may be most effective when paired with other targeted interventions or in settings where broad behavioral change is achievable and well-aligned with transmission pathways.

#### County Demographics Do Not Reliably Predict NPI Effectiveness

To assess whether basic county-level demographic characteristics could explain the variation in intervention effectiveness observed across counties, we compared the median reduction in peak hospitalizations for each intervention to a demographic feature related to that intervention (figure 5). For example, school closure effects were compared to the proportion of children under 18, workplace closure effects were compared to the labor force participation rate.

**Figure 5:**
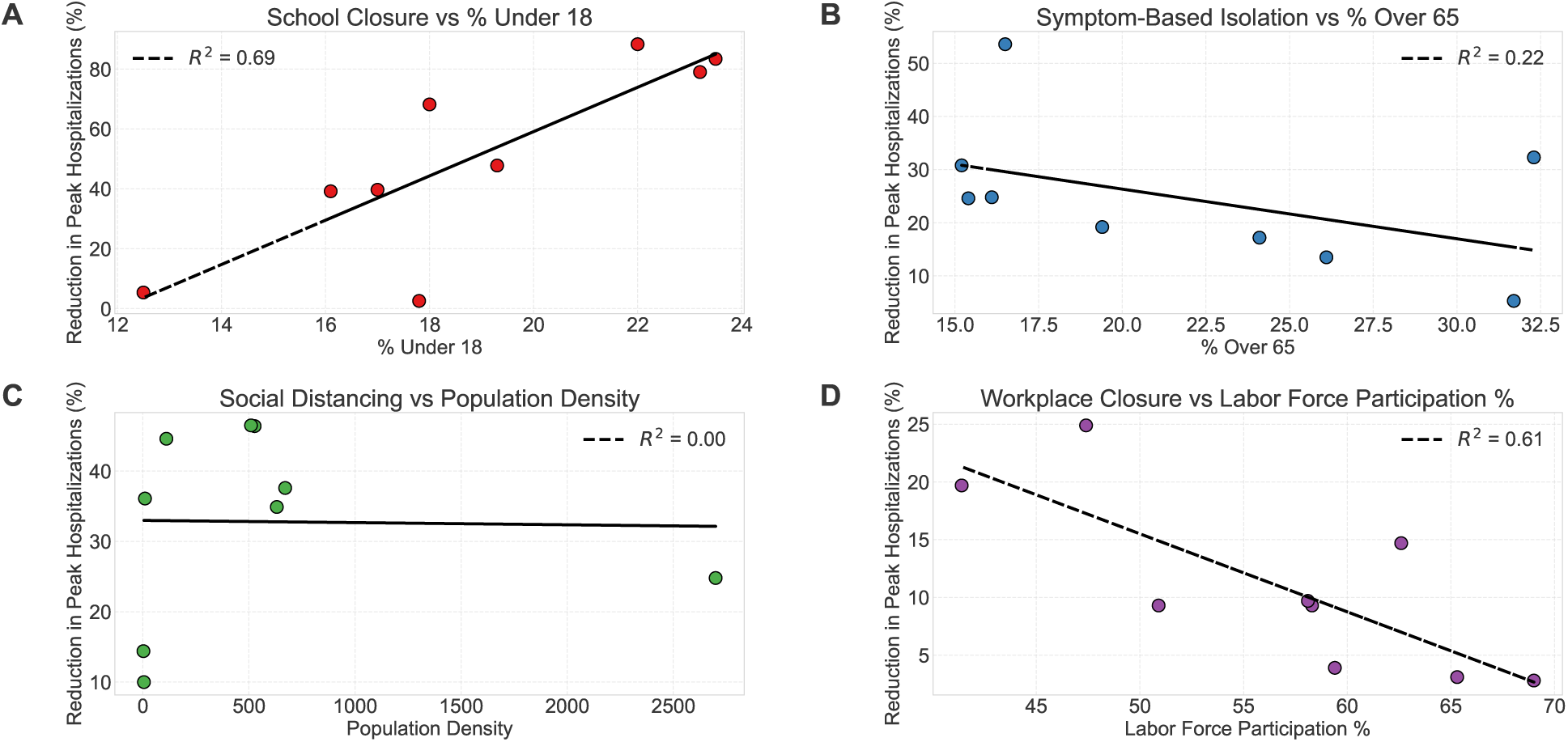
County demographics only predict efficacy of some interventions. Each panel shows the relationship between the median effect of an intervention and a related demographic characteristic across Michigan counties. **(A)** School closure efficacy was positively associated with the percentage of children under 18 (*R*^2^ = 0.69). **(B)** Symptom-based isolation showed a weak negative trend with the proportion of adults over 65 (*R*^2^ = 0.22). **(C)** Social distancing effectiveness showed no relationship with population density (*R*^2^ = 0.00). **(D)** Workplace closure effectiveness showed a surprising negative association with labor force participation rate (*R*^2^ = 0.61), likely reflecting confounding structural factors rather than a causal relationship.

Among the four interventions examined, only school closures and workplace closures showed even moderate associations with their corresponding demographic predictors (child population share and labor force participation rate). School closures were more effective in counties with a higher proportion of children under 18 (*R*^2^ = 0.69), a trend that is both intuitive and expected given the role of schools in facilitating youth transmission. Workplace closures, however, showed a surprising negative association with labor force participation rate (*R*^2^ = 0.61), suggesting they were more effective in counties where fewer working-age adults were employed. This is unlikely to be a causal relationship; more plausibly, it reflects that workplace closure efficacy varies due to unmeasured structural or behavioral features not captured by basic demographics.

In contrast, the other interventions showed weak or no relationship with the chosen demographic predictors. Symptom-based isolation showed a slight negative trend with the proportion of residents over 65 (*R*^2^ = 0.22), and social distancing showed no relationship at all with population density (*R*^2^ = 0.00), despite expectations that higher-density areas might benefit more from reduced contacts.

Together, these findings underscore that county demographics alone are poor predictors of which interventions will be most effective in a given setting. Instead, they highlight the importance of a simulation-based approach that captures dynamic interactions between demographic structure, contact patterns, and intervention mechanisms under uncertainty, enabling more accurate and context-specific policy guidance.

### Sensitivity of Intervention Efficacy to Pathogen Biology

One goal of our framework is to support prospective use in some situations. For example, during the COVID-19 pandemic, there were multiple successive waves. Thus, a reasonable use of our approach may be to use it retrospectively to estimate intervention efficacy on the first wave and then apply the best performing intervention on subsequent waves. For this strategy to be effective, it must be the case that the efficacy of interventions does not meaningfully change from wave to wave. We hypothesize that this is true, and that intervention efficacy is more dependent on the (relatively) fixed contact structures of the county than the variations in pathogen biology from one wave to the next. In this section, we evaluate the sensitivity of intervention efficacy to changes in pathogen biology to answer this question.

Specifically, we consider two questions:

1. When parameters are perturbed as they would be during a wave change, what percent of the time does the most effective intervention stay the same?
2. Under the same perturbation, how much does intervention efficacy change?

To test this, we generated 250 biological parameter sets via Latin Hypercube Sampling over the ranges in figure 2 for transmissibility, asymptomatic transmissibility, latent period, hospitalization probability, and hospitalization duration. For each parameter set, we ran one baseline simulation and one for each of our candidate interventions (across 5 random seeds) and measured the reduction in peak hospitalizations in Washtenaw county.

First, to assess how stable the rankings of intervention efficacy were, we computed pairwise win rates for each intervention pair (the percentage of the time intervention A is more effective at reducing peak hospitalizations than intervention B across parameters and seeds). Then, to estimate the marginal sensitivity of interventions to pathogen biology (i.e., calculate the expected change in efficacy for wave-to-wave pathogen shift), we fit least squares regressions of efficacy to the five biological parameters for each intervention. We then multiplied the regression coefficients by plausible perturbation magnitudes to produce expected shifts in efficacy in absolute percentage points.

We found that the most effective intervention was stable. In Washtenaw county, across 91.8% of runs, the most effective intervention was school closure. The second most effective was more variable, since stay-home-if-symptomatic and social distancing interventions already had similar efficacy.

For marginal sensitivity, a 0.1-unit increase in transmissibility (similar to the initial difference between counties) shifted efficacy by roughly 5–6 percentage points for school closure, but less for other interventions. Increasing asymptomatic transmissibility made all interventions more effective (most likely because the disease was more severe) except stay-home if symptomatic, which is predictable. Hospitalization probability and duration had little effect across interventions, probably because contact-based interventions reduce exposures upstream of clinical outcomes. Table 1 shows the change in intervention efficacy observed when perturbing the biological parameters.

**Table 1:** Expected change in peak hospitalization reduction (percentage points) per biological perturbation. Values are least squares coefficients scaled by Δ.

| **Intervention** | $\Delta$ Trans.<br>= 0.10 | $\Delta$ Asymp. Trans.<br>= 0.10 | $\Delta$ Latent<br>= 1 d | $\Delta$ Hosp. Prob.<br>= 0.02 | $\Delta$ Hosp. Length<br>= 2 d |
| --- | --- | --- | --- | --- | --- |
| School closure | +5.8 | +3.4 | -1.8 | -0.1 | -0.7 |
| Social distancing | +2.3 | +1.6 | -1.2 | +0.04 | -0.4 |
| Stay-home if symptomatic | +2.0 | -1.8 | -1.0 | +0.03 | -0.5 |
| Workplace closure | +0.8 | +0.9 | -0.02 | +0.06 | +0.01 |

Overall, the most effective intervention was fairly stable, staying the same for 91.8% of simulations we ran. In addition, the changes in efficacy were small when biological parameters were perturbed. Only one intervention-parameter combination had an average change in efficacy greater than 5% (5.8%), and 11/20 of the average changes were less than 1%.

## 3 Discussion

We present a framework that couples agent-based simulation with surrogate-informed inference to evaluate non-pharmaceutical interventions at the county level. The most important feature of the approach is that it does not commit to a single best-fit parameter set. Routine case and hospitalization data are not sufficient to pin down individual ABM parameters, and many combinations of transmissibility, latent period, asymptomatic fraction, reporting rate, and severity can reproduce the same observed trajectories. Rather than selecting one such combination, the framework retains the full set of ABM parameter combinations consistent with the data and propagates this set forward into intervention simulations. The result is a distribution of intervention outcomes that reflects the uncertainty the data leave unresolved. Interventions whose effects are stable across this distribution can be acted on with greater confidence. Interventions whose effects vary substantially indicate that the available data leave the predicted impact ambiguous.

Applied to COVID-19 in nine demographically distinct Michigan counties, the framework produces clear differences in the ranking and robustness of four non-pharmaceutical interventions. Under the intervention implementations used here, school closures produced large and stable reductions in peak hospitalizations in counties with substantial K-12 populations and limited reductions in older rural counties. Symptom-based isolation reduced peak burden in many parameter regimes but was sensitive to the assumed asymptomatic fraction. Workplace closures produced modest and inconsistent effects across counties. General social distancing produced moderate but parameter-sensitive effects. The specific magnitudes reported in the results section depend on these implementation assumptions. Alternative compliance, contact-reduction, or behavioral assumptions can be substituted into the framework without changes to the inference pipeline. We also found that demographics cannot reliably forecast which interventions will succeed, motivating our approach of using locally fit mechanistic simulations for intervention evaluation.

The most natural use case for the framework is between-wave preparedness for diseases with recurring waves, where surveillance data from one wave can be used to characterize the data-consistent parameter space for a region, and candidate interventions can then be evaluated in advance of the next wave. The validity of this use depends on whether conclusions garnered from one wave transfer to the next. We have two pieces of evidence that they do. The temporal hold-out validation in the appendix shows that calibrating the framework on the first 80 days of the wave and projecting forward over the remaining 40 days produces ABM trajectories that track the held-out data, with parameter ranges that change modestly under the reduced training window, demonstrating that there is stability over time in the model fits and thus conclusions. Also, sensitivity analysis in which biological parameters (transmissibility, latent period, asymptomatic fraction, hospitalization probability and duration) are perturbed while contact structure is held fixed (we would not expect contact structure to vary wave-to-wave) shows that the most effective intervention within a county is the same across 91.8% of runs with varying parameter biology in section 4. The intuition is that NPI effectiveness depends primarily on the contact structure that each intervention targets, which is a property of the local population that is stable on the timescale of months to years, rather than on pathogen biology, which can shift across waves. Together these analyses support the use of the framework for between-wave planning even when pathogen characteristics shift.

A key methodological limitation is the potential for surrogate model misspecification. The surrogate is a deliberate simplification of the ABM. If it omits a mechanism that meaningfully shapes the dynamics, the mapping from ABM parameter space to observed data may be systematically biased. For example, if the surrogate model lacked underreporting parameters, which were critical for COVID-19, the framework might struggle to correctly map ABM parameter space to observed data [16]. The extent to which the high-dimensional ABM parameter space can compensate for structural omissions in the surrogate is an open question. In some cases, the flexibility of the ABM may allow it to recover appropriate dynamics despite surrogate misspecification. In others, missing mechanisms in the surrogate could systematically bias the identified ABM parameter region. Understanding the robustness of this approach to different forms and degrees of surrogate misspecification is an important direction for future work, and may inform best practices for surrogate model selection in different outbreak contexts.

Our framework operates on standard surveillance data, so it can be applied to any pathogen (although the FRED model right now only has local populations available in the US and India [13]). It complements national-scale forecasting efforts by supporting localized intervention analysis tailored to the demographic and infrastructural characteristics of individual communities.

## 4 Methods

### Agent-Based Simulations

We used the FRED (Framework for Reconstructing Epidemiological Dynamics) platform to simulate COVID-19 outbreaks under varying parameterizations [13]. Parameter sets were generated using Latin Hypercube Sampling (LHS) across ranges for latent period, transmissibility (overall and asymptomatic), hospitalization probability and duration, and case and hospitalization reporting rates. Each parameter set was run with five stochastic replicates to capture variability. Model outputs included daily symptomatic cases and hospitalizations, which were aggregated and passed through the same surrogate fitting procedure as the observed surveillance data to ensure consistency.

### Surrogate Model Selection and Fitting

To link ABM simulations with observed epidemic trajectories, we evaluated five candidate ODE-based surrogate models (SIRH, SEIRH, SEAIRH, SEAI_1_I_2_RH, SEIRHD), each of which had parameters governing reporting rates for both cases and hospitalizations. Models were fit to county-level COVID-19 case and hospitalization time series [14] using weighted least squares, and compared using the Akaike Information Criterion (AIC) [17]. The SIRH model provided the best overall fit in all three main representative counties (ΔAIC = 24.29 in Washtenaw, 2.99 in Wayne, and 31.91 in Ontonagon). Based on these results, SIRH was selected as the surrogate for all downstream analyses. Parameter confidence intervals were estimated using the profile likelihood method.

### Parameter Surface Reconstruction

Within the SMoRe ParS framework, we mapped ABM parameters to surrogate model (SM) parameters to quantify uncertainty and constrain mechanistic space. For each ABM configuration, we ran simulations, fit the surrogate, and estimated confidence intervals using profile likelihood. These ABM–SM mappings were interpolated to form continuous response surfaces, which were then compared with surrogate fits to real data. ABM parameter sets consistent with the empirical 95% confidence regions were retained, defining county-specific parameter spaces for downstream intervention analyses.

### Data Sources and Calibration

We obtained COVID-19 case and hospitalization data from public health surveillance systems at the county level in Michigan [14]. Case data consisted of daily incidence counts, while hospitalization data represented total hospital occupancy (i.e., prevalent, not incident, hospitalizations). We used census data for population [18]. No preprocessing or smoothing was applied to the data. However, due to known underreporting of COVID-19 cases and hospitalizations, we fit an underreporting rate as an additional parameter in the surrogate model calibration process.

### Intervention Simulation and Robustness Evaluation

To assess how parameter uncertainty affects the projected outcomes of public health interventions, we simulated several non-pharmaceutical interventions within the FRED platform across the full range of parameter combinations consistent with observed data. The interventions evaluated included: (1) school closures (implemented by eliminating school and classroom contact rates), (2) stay-home-if-symptomatic policies (increasing probability of staying home to 60% [19], (3) non-essential work closures (implemented by reducing workplace and office contact rates by 66% [20]), and (4) general social distancing (implemented by reducing social contact rates by 60% [21]).

For each intervention, we evaluated outcomes using three primary metrics: (1) peak daily cases, (2) peak hospitalizations, and (3) cumulative burden of disease (total cases and total hospitalizations). Intervention robustness was assessed by evaluating performance at multiple locations within the identified ABM parameter region for each county. Interventions that performed consistently well across this space were classified as robust, while those whose effectiveness varied substantially were deemed sensitive to parameter uncertainty.

### Implementation

All surrogate model fitting, profile likelihood computation, and intervention evaluation were implemented in Python. Agent-based simulations were run using the FRED simulation platform with custom scenario definitions. All code used in this study, including scripts for surrogate model fitting, ABM execution, parameter surface reconstruction, and intervention analysis, is available at https://github.com/carsondudley1/smoreparscovid.

## Data Availability

All data used in this study are available via MI Start Map (https://mistartmap.info/active-metrics.html). The simulation framework, FRED, is available at https://fred.publichealth.pitt.edu/.

## 5 Author Contributions

C.D. led the study and implemented the code. D.B., H.J., K.-A.N., and E.R. contributed equally to this work. D.B. created the conceptual figure. K.-A.N. performed the statistical significance analyses. M.E. provided guidance on relevant public health questions and framing. C.D., H.J., and T.J. planned the project. All authors shaped the direction of the project and reviewed and edited the manuscript.

## Appendices

### Agent-Based Model Parameter Sampling

We used Latin Hypercube Sampling (LHS) to generate parameter sets for agent-based simulations across the full range of plausible epidemiological values. The same parameter ranges and sampling strategy were applied to all three counties, with the same sampled parameters used across counties. Seven key parameters were varied, with the following continuous sampling ranges:

**Table 2:**
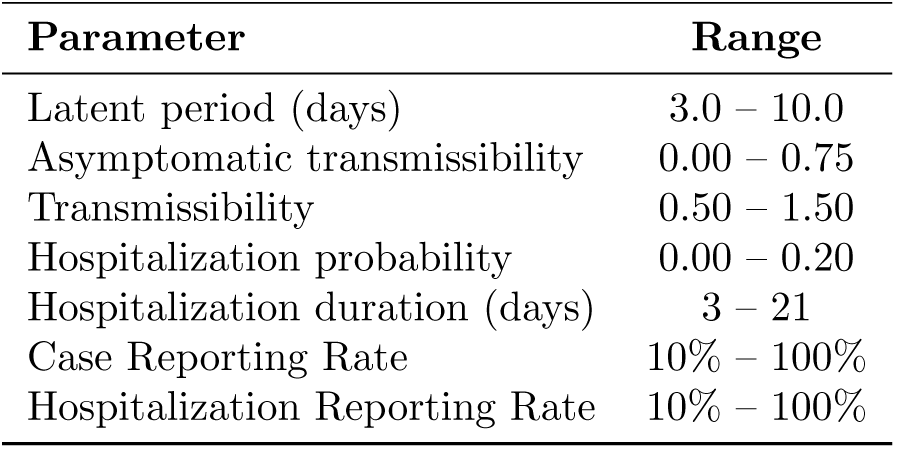
Parameter ranges used for ABM sampling.

| Parameter | Range |
| --- | --- |
| Latent period (days) | 3.0 – 10.0 |
| Asymptomatic transmissibility | 0.00 – 0.75 |
| Transmissibility | 0.50 – 1.50 |
| Hospitalization probability | 0.00 – 0.20 |
| Hospitalization duration (days) | 3 – 21 |
| Case Reporting Rate | 10% – 100% |
| Hospitalization Reporting Rate | 10% – 100% |

A total of 250 unique parameter combinations were generated using Latin Hypercube Sampling, with each parameter sampled continuously and independently within its specified range. For each parameter set, 5 independent stochastic replicates were simulated to mitigate random variability arising from the agent-based dynamics. All simulations were initialized identically across replicates to ensure comparability.

### Intervention Implementation Details

All interventions were simulated under consistent compliance assumptions across counties. Because the study period captured an ongoing epidemic wave rather than epidemic initiation, simulations were initialized with seeded infections and hospitalizations based on observed surveillance data for each county as of August 28, 2020. Interventions were applied from the beginning of each simulation and remained in effect throughout the full simulation period. Specific implementation assumptions were as follows: school closures were modeled by fully eliminating school and classroom contact rates; stay-home-if-symptomatic policies increased the daily probability of isolation for symptomatic individuals to 60% [19]; non-essential work closures were modeled by reducing workplace and office contact rates by 66% [20]; and general social distancing was modeled by reducing community and social contact rates by 60% [21].

### Surrogate Model Fitting

To connect agent-based model (ABM) simulations to observed epidemic trajectories, we evaluated several compartmental ordinary differential equation (ODE) models as candidate surrogates. The models considered were:

- SIRH (Susceptible–Infectious–Recovered–Hospitalized):

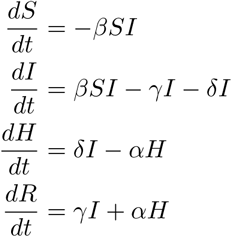
- SEIRH (Susceptible–Exposed–Infectious–Recovered–Hospitalized):

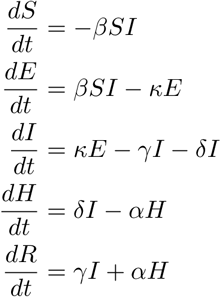
- SEAIRH (Susceptible–Exposed–Asymptomatic–Infectious–Recovered–Hospitalized):

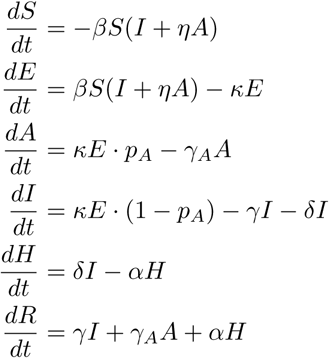
- SEAI_1_I_2_RH (with separate compartments for mild and severe infection):

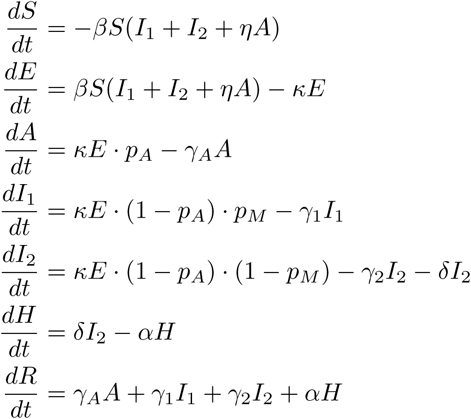
- SEIRHD (Susceptible–Exposed–Infectious–Recovered–Hospitalized–Deceased):

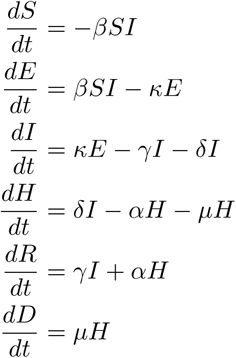

Each model was fit to observed COVID-19 case and hospitalization time series using a weighted least squares approach. Model selection was performed using the Akaike Information Criterion (AIC) [17]. The SIRH model provided the best fit across all counties and was therefore selected as the final surrogate model for all downstream analysis.

Model fitting was performed via weighted least squares under an observation model incorporating both underreporting and measurement error. Specifically, we estimated eight parameters jointly:

- *β*: transmission rate
- *γ*: recovery rate
- *δ*: hospitalization rate
- *α*: hospital discharge rate
- *ρ*_cases_: reporting rate for cases
- *ρ*_hosp_: reporting rate for hospitalizations
- *σ*_cases_: observation noise for cases
- *σ*_hosp_: observation noise for hospitalizations

Model fitting was performed via weighted least squares. The objective function minimized was:

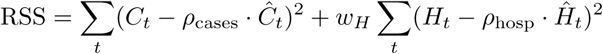

where *C_t_* and *H_t_* are the observed daily cases and hospitalizations at time *t*, *Ĉ_t_* and *Ĥ_t_* are model-predicted quantities, and *w_H_* is a weighting factor applied to hospitalization residuals to ensure both data streams contribute meaningfully to the fit despite differences in scale. We used *w_H_* = 5.

Optimization was performed using the L-BFGS-B algorithm with multiple random initializations to avoid local minima. Parameter bounds were specified for each parameter (see Table 3). The full epidemic time series for each county was fit jointly to both case and hospitalization data.

**Table 3:**
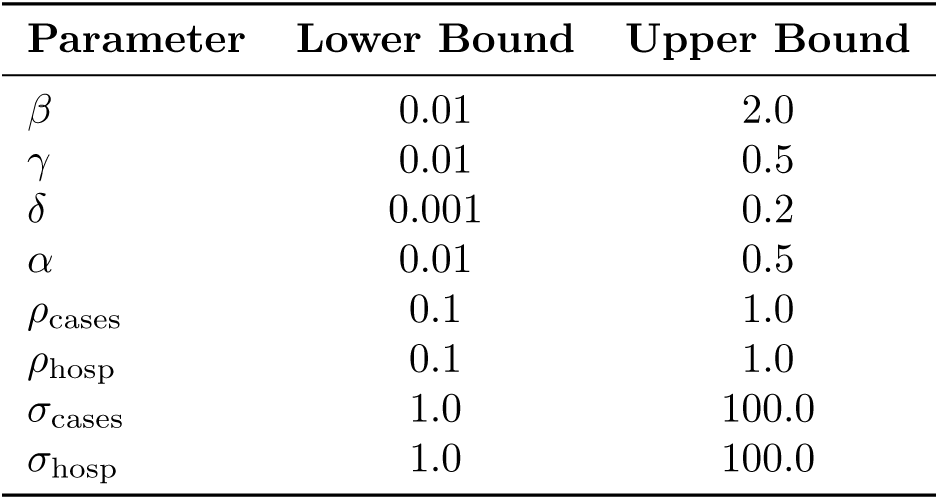
Parameter bounds used in surrogate model optimization.

| Parameter | Lower Bound | Upper Bound |
| --- | --- | --- |
| $\beta$ | 0.01 | 2.0 |
| $\gamma$ | 0.01 | 0.5 |
| $\delta$ | 0.001 | 0.2 |
| $\alpha$ | 0.01 | 0.5 |
| $\rho_{\text{cases}}$ | 0.1 | 1.0 |
| $\rho_{\text{hosp}}$ | 0.1 | 1.0 |
| $\sigma_{\text{cases}}$ | 1.0 | 100.0 |
| $\sigma_{\text{hosp}}$ | 1.0 | 100.0 |

#### Parameter Surface Reconstruction

Following the SMoRe ParS framework, we constructed explicit mappings between ABM parameters and surrogate model (SM) parameters to enable systematic uncertainty quantification and data-informed constraint of mechanistic model space. For each ABM parameter combination sampled via Latin Hypercube Sampling, we simulated the agent-based model, extracted summary trajectories (e.g., hospitalization time series), and fit the SIRH surrogate model to these trajectories using maximum likelihood estimation.

This procedure yielded surrogate parameter estimates for each sampled ABM parameter combination. To quantify uncertainty in these estimates, we applied the profile likelihood method to each ABM–SM fit, calculating 95% confidence intervals for each surrogate parameter by profiling the objective function with respect to each parameter while optimizing over the others. This yielded, at each ABM parameter combination, confidence intervals on the surrogate parameters conditional on that ABM setting.

The resulting discrete mappings—from ABM parameter vectors 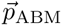 to surrogate parameter estimates and their confidence intervals—were then used to reconstruct continuous response surfaces via multivariate interpolation. Specifically, we used `scipy`’s `LinearNDInterpolator` to construct interpolated functions of the form:

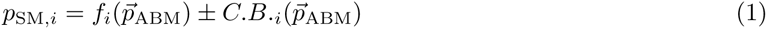

where *p*_SM*,i*_ denotes the *i*th surrogate parameter (e.g., *β, γ, δ, α*), *f_i_* is the interpolated best-fit value, and C.B.*_i_* represents the corresponding 95% confidence bounds, both obtained via interpolation from the set of profile-likelihood-derived estimates.

To determine which ABM parameter combinations are consistent with the real-world data, we first fit the surrogate model directly to the observed data using weighted least squares and profile likelihood, generating a data-derived confidence region for each surrogate parameter. We then used the reconstructed response surfaces to project every ABM parameter vector 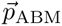 into surrogate space, and retained only those ABM parameter combinations whose projected surrogate values fell within the empirical 95% confidence region inferred from data. The union of these retained points defines an implicit confidence region over ABM parameters, incorporating both data-derived surrogate uncertainty and interpolation-based ABM–surrogate relationships.

This reconstruction process was performed independently for each county, enabling region-specific parameter space characterization while maintaining a consistent calibration pipeline across all study sites. The resulting confidence regions over ABM parameters support downstream analyses such as policy ranking, robustness checks, and posterior predictive simulations within data-consistent mechanistic regimes.

#### Results for Additional Counties

To evaluate whether the intervention patterns observed in Washtenaw, Wayne, and Ontonagon Counties held across structurally similar settings, we analyzed six additional Michigan counties grouped by demographic and infrastructural similarity. Specifically, we selected:

- **Like Washtenaw (younger, university-centered):** Isabella County and Ingham County (also contains state capital)
- **Like Wayne (urban, high comorbidity burden):** Genesee County and Kent County
- **Like Ontonagon (rural, aging population):** Luce County and Iron County

**Table 4:** ABM Parameter estimates recovered by the SMoRe ParS framework for the additional counties. Values represent point estimates with 95% confidence intervals derived from model calibration to case and hospitalization data.

| ABM Parameter | Isabella | Ingham | Genesee | Kent | Luce | Iron |
| --- | --- | --- | --- | --- | --- | --- |
| Hospitalization Probability | 5.9%<br>[3.3, 7.9] | 6.2%<br>[5.1, 7.6] | 8.2%<br>[6.8, 12.5] | 7.2%<br>[5.8, 10.3] | 8.2%<br>[6.9, 10.1] | 9.2%<br>[8.6, 10.1] |
| Hospitalization Duration (days) | 6.4 [4.5, 14.0] | 8.6 [3.5, 12.1] | 11.9 [11.2, 12.4] | 9.9 [8.5, 14.0] | 7.9 [6.5, 9.0] | 10.1 [8.8, 14.0] |
| Latent Period (days) | 5.2<br>[5.02, 5.40] | 4.65<br>[4.4, 4.89] | 5.16<br>[4.84, 5.63] | 4.76<br>[4.54, 5.01] | 3.76<br>[3.54, 3.98] | 5.98<br>[5.39, 7.01] |
| Transmissibility | 1.63<br>[1.54, 1.77] | 1.81<br>[1.68, 1.98] | 1.90<br>[1.83, 1.98] | 1.78<br>[1.73, 1.82] | 1.76<br>[1.71, 1.80] | 1.49<br>[1.43, 1.60] |
| Asymptomatic Transmissibility | 36.0%<br>[35.1, 38.5] | 37.3%<br>[36.7, 39.6] | 38.3%<br>[37.1, 39.8] | 37.3%<br>[37.1, 37.8] | 38.3%<br>[37.6, 38.9] | 37.6%<br>[37.3, 38.0] |
| Case Reporting Rate | 14.0%<br>[11.3, 16.4] | 16.1%<br>[12.4, 19.9] | 26.3%<br>[21.2, 30.2] | 22.0%<br>[18.2, 26.2] | 6.0%<br>[6.0, 12.3] | 24.2%<br>[18.0, 27.3] |
| Hospitalization Reporting Rate | 93.0%<br>[86.5, 100.0] | 100.0%<br>[93.2, 100.0] | 97%<br>[91.1, 100.0] | 100.0%<br>[95.1, 100.0] | 6.0%<br>[6.0, 10.1] | 11.0%<br>[10.0, 19.1] |

**Figure 6:**
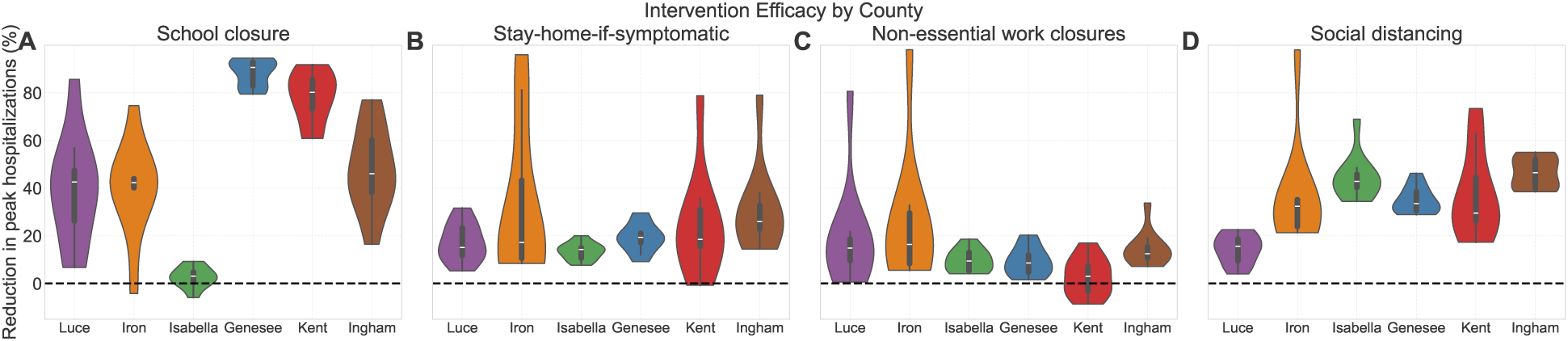
Intervention effectiveness across additional counties. Violin plots show the distribution of reductions in peak hospitalizations (%) under four non-pharmaceutical interventions, simulated across data-consistent ABM parameter combinations.

**(A) School closures** remained highly effective in counties structurally similar to Washtenaw and Wayne. Ingham, Kent, and Genesee exhibited the largest reductions in peak hospitalizations. Ingham showed strong but slightly more variable reductions. An exception was Isabella County, where school closures produced minimal effect. This likely reflects the unique population structure of Isabella, which has a disproportionately high university student population and relatively fewer school-aged children. Because our intervention model targets K–12 settings and does not modify university transmission, the impact of school closures is possibly muted in counties where universities dominate the educational landscape. In contrast, rural Ontonagon-like counties (Luce and Iron) also showed impactful but highly variable school closure effects.
**(B) Stay-home-if-symptomatic** policies again showed the strongest performance in university-oriented counties. Ingham and Isabella mirrored Washtenaw’s high median reductions, while Wayne-like counties (Genesee, Kent) had more moderate effects. Ontonagon-like counties (Luce, Iron) showed wide variance and relatively weak performance, reinforcing the policy’s vulnerability to asymptomatic transmission and limited control in sparse rural networks.
**(C) Non-essential work closures** continued to produce modest and inconsistent reductions across all counties, with little structural variation. Even in more urban counties like Genesee and Kent, median effects were small, and wide spreads were sometimes observed, particularly in Iron and Luce. These findings support the main conclusion that work closures are a low-impact strategy in most contexts.
**(D) Social distancing** remained moderately effective in urban and university-adjacent counties. Isabella, Genesee, and Ingham saw stable, substantial reductions. In Kent and Iron, the results were also positive but more variable. In Luce, the results were much less substantial, possibly reflecting lower baseline connectivity due to rurality. Isabella and Ingham had the highest average effect, possibly reflecting highest baseline connectivity due to the university settings.

Overall, these results reinforce the main text conclusions:

- **School closures** are consistently robust in populations with many school-aged children.
- **Symptom-based isolation** is highly context-dependent and weakened by asymptomatic spread.
- **Work closures** produce only modest effects.
- **Social distancing** can produce strong effects particularly in high connectivity settings.

These additional counties support the conclusion that intervention performance is tightly coupled to local demographic and structural features, and that effective outbreak policy design must account for these context-specific factors.

#### Out-of-Sample Validation

To assess the reliability and stability of our framework, we performed a temporal hold-out validation. We calibrated the agent-based model (ABM) using only the first 80 days of the epidemic wave for Wayne and Washtenaw counties—a period primarily characterized by exponential growth—and evaluated the resulting simulations against the full 120-day observed time series.

In Wayne County, where the 80-day training window included the initial peak, the resulting ABM projections remained highly consistent with the held-out data (figure 7). The parameters identified using the partial dataset did not deviate significantly from those found using the full 120-day window, yielding a stable fit across both periods.

For Washtenaw County, the validation provided a more rigorous test as the outbreak had not yet peaked by day 80 and was still in the exponential growth phase. Calibrating solely on this growth period resulted in a higher transmissibility estimate compared to the full-data fit (point estimate 1.92 vs. 1.73). Interestingly, while the surrogate model had an accurate fit to the 80-day training period (which is expected, because it was fit on that), the resulting ABM projection exhibited a slight overshoot during the initial training period rather than the held-out window (figure 7). For the surrogate model, the higher estimated transmissibility led it to peak later, with a good fit initially and then an overshoot. For the ABM, the peak timing was more accurate, but the initial growth phase was overshot. These results indicate that, while fitting to a narrow window of exponential growth can introduce bias in parameter fitting, especially for our simplified surrogate models, our method’s projection into the higher-dimensional ABM space potentially provides a corrective mechanism. By connecting the data-consistent parameter sets to a realistic population architecture, the framework mitigates the impact of incomplete surveillance signals, yielding fits that are more representative of the full epidemic wave and more biologically plausible than would be expected from the limited training window.

To be clear, our goal is validation of the structural stability of our approach, not proposing use for real-time epidemic forecasting. While forecasting models optimize predictive accuracy (sometimes with mechanistic models similar to what we use but often with statistical or deep learning approaches [22, 23, 24, 25]), the goal of our framework is to identify data-consistent parameter regimes that allow for robust policy evaluation. Our hold-out analysis demonstrates that even when calibrated on incomplete epidemic phases, the mapping to the higher-dimensional ABM structure provides some regularization that prevents the model from drifting into potentially biologically implausible regimes (like extreme transmission).

#### Statistical Analysis

##### Outcome

All tests use the ratio of peak hospitalizations under an intervention to the baseline (no intervention) peak for the same county and parameter setting. Ratios *<* 1 indicate reductions; ratios > 1 indicate increases.

##### Design and error control

For each comparison family we ran a one-way ANOVA and, when significant, Tukey’s HSD for all pairwise contrasts (familywise error control within the family). We report *p*-values; distributional summaries are shown in the main-text figures.

#### Across counties (per intervention)

For each intervention, we compared distributions across Washtenaw, Wayne, and Ontonagon (each *n* = 30).

- **School closure.** ANOVA *p* = 7.04 × 10*^−^*^58^; all pairwise contrasts significant (Tukey *p <* 10*^−^*^10^).
- **Social distancing.** ANOVA *p* = 1.17×10*^−^*^18^; all pairwise contrasts significant (Tukey *p* ≤ 2.6×10*^−^*^5^).
- **Workplace closure.** ANOVA *p* = 0.312; no pairwise differences significant (all Tukey *p* ≥ 0.338).
- **Stay-home-if-symptomatic.** ANOVA *p* = 6.50 × 10*^−^*^26^; all pairwise contrasts significant (Tukey *p* ≤ 7.5 × 10*^−^*^6^).

#### Within counties (across interventions)

For each county, we compared school closure, social distancing, workplace closure, and stay-home-if-symptomatic (each *n* = 30).

- **Wayne County.** ANOVA *p* = 2.49 × 10*^−^*^104^; all pairwise contrasts significant (smallest *p* = 0.003 for social vs. symptom).
- **Washtenaw County.** ANOVA *p* = 4.64 × 10*^−^*^29^; all pairwise contrasts significant except social vs. symptom (Tukey *p* = 0.247, n.s.).
- **Ontonagon County.** ANOVA *p* = 0.387; no pairwise differences significant (all Tukey *p* ≥ 0.323).

**Figure 7:**
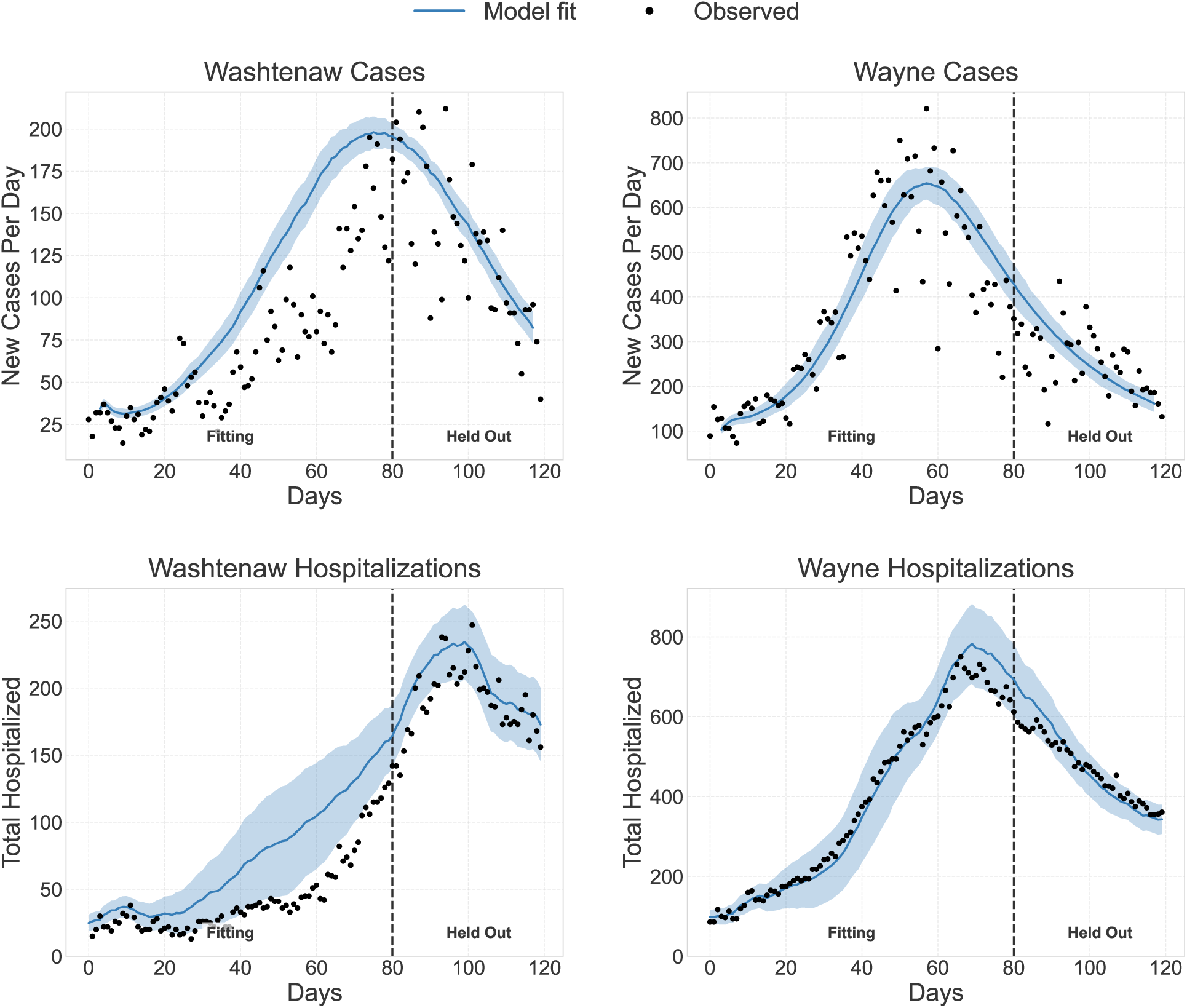
Out-of-Sample Validation in Wayne and Washtenaw Counties. The framework was calibrated using surveillance data from the first 80 days (fitting) and projected forward for the remaining 40 days (which were held out from fitting). Solid lines represent the ABM mean across 10 simulations, with shaded regions indicating ±1 SD across stochastic replicates. In Wayne County, parameters remained stable, yielding an accurate fit across both windows. In Washtenaw County, calibrating solely on the exponential growth phase (pre-peak) resulted in higher transmissibility estimates, leading to a slight overshoot during the initial period, though the model relatively successfully anticipated the peak timing and subsequent decay rate.

##### Interpretation

Across counties, school closure, social distancing, and symptom-based isolation show statistically detectable differences in effect, whereas workplace closure does not. Within counties, intervention effects differ in Wayne and Washtenaw but are not statistically distinguishable in Ontonagon, consistent with greater stochastic variability in small, sparse populations.

##### Assumptions and robustness

ANOVA and Tukey’s HSD assume approximate normality and homoscedasticity. Given bounded ratios and potential heteroskedasticity, we emphasize robust summaries (medians/IQRs) in the figures. As a sensitivity check, results were qualitatively unchanged under nonparametric testing (Kruskal–Wallis with Dunn’s post hoc and Holm correction), and under percentile bootstrap confidence intervals for median differences (not shown).

#### ABM Parameter Estimates and Comparison to Empirical Data

For each county, we inferred the subset of ABM parameter combinations whose surrogate model projections matched the observed case and hospitalization time series. This calibration step yielded joint estimates of epidemiological parameters, including transmissibility, latent period, asymptomatic fraction, hospitalization risk and duration, and reporting rates. Confidence intervals were derived by selecting ABM parameters whose surrogate projections fell within the 95% bounds of the fitted surrogate model (see Methods: Parameter Surface Reconstruction). These data-consistent parameter regions are the inputs to the intervention analyses in the main text. Here we summarize the marginal distributions of each parameter for the three representative counties and compare them to independent empirical estimates.

The SMoRe ParS framework identified county-specific parameter regions whose marginal summaries are broadly compatible with the literature (Table 5). Hospitalization probabilities follow the expected demographic gradient: Washtenaw County had the lowest estimate at 4.9%, consistent with its younger population. Wayne County was higher at 7.0%, consistent with a slightly older population and higher chronic disease burden, and Ontonagon County had the highest at 9.2%, consistent with its older age distribution. These estimates are in line with age-stratified severity estimates established early in the pandemic [26]. The recovered latent periods (3.7–5.85 days) span the range reported in clinical studies of the SARS-CoV-2 incubation period [27, 28]. Asymptomatic transmissibility was consistent across counties (37.5–38.5%), which is the expected behavior if asymptomatic infectiousness is governed primarily by pathogen biology rather than local demographic composition.

The reporting rates vary across counties in ways that are also interpretable. Wayne County’s lower case reporting rate (9.1%) is consistent with reports of strained testing capacity in dense urban settings during this period. Ontonagon County’s low hospitalization reporting rate (10%) likely reflects the absence of a full-service hospital in the county: severe cases were transported to neighboring counties and would not appear in county-level hospitalization counts. Jointly fitting both data streams and estimating reporting rates separately allowed the framework to recover relatively plausible hospitalization risk estimates even in this sparse-data setting.

Some recovered parameters vary across counties in ways that do not have a clear biological or demographic interpretation. The latent period estimate for Wayne County (5.85 days) is at the upper end of the clinical range and substantially longer than for Washtenaw (3.7 days), despite no obvious reason the underlying biology of SARS-CoV-2 should differ between adjacent populations. The hospitalization duration estimates show wide confidence intervals (Washtenaw: 3.5–14.0 days; Wayne: 3.5–12.1 days), reflecting limited identifiability from hospitalization count data alone. These features illustrate the broader point that motivates the main-text analyses: marginal point estimates from a single calibration are an incomplete summary of the data-consistent parameter region. The joint distribution over ABM parameters that the framework retains can support an observed trajectory through many different combinations of transmission, severity, and reporting, and the appropriate use of these inferred regions is to propagate them forward into intervention evaluation rather than to read them as highly accurate point estimates of biological quantities.

**Table 5:** ABM Parameter estimates recovered by the SMoRe ParS framework for Washtenaw, Wayne, and Ontonagon Counties. Values represent point estimates with 95% confidence intervals derived from model calibration to case and hospitalization data.

| **ABM Parameter** | **Washtenaw** | **Wayne** | **Ontonagon** |
| --- | --- | --- | --- |
| Hospitalization Probability | 4.9% [3.3, 5.9] | 7.0% [6.6, 9.4] | 9.2% [3.8, 9.2] |
| Hospitalization Duration (days) | 5.4 [3.5, 14.0] | 8.6 [3.5, 12.1] | 10.9 [10.5, 11.3] |
| Latent Period (days) | 3.7 [3.6, 4.0] | 5.85 [5.6, 6.02] | 4.22 [3.87, 4.53] |
| Transmissibility | 1.73 [1.64, 1.87] | 1.81 [1.68, 1.98] | 1.50 [1.43, 1.65] |
| Asymptomatic Transmissibility | 38.0% [37.1, 42.3] | 37.5% [36.9, 39.6] | 38.5% [37.5, 39.8] |
| Case Reporting Rate | 14.0% [10.1, 17.1] | 9.1% [5.9, 13.9] | 26.3% [21.2, 30.2] |
| Hospitalization Reporting Rate | 82.0% [75.3, 84.2] | 71.3% [65.3, 78.1] | 10.0% [10.0, 11.3] |

